# Integrated genomic meta-analysis identifies cell-type-specific signatures and targets in early Alzheimer’s disease

**DOI:** 10.64898/2026.09.21.26362150

**Authors:** Moritz Steinruecke, J Kenneth Baillie, Zoeb Jiwaji

**Affiliations:** University of Cambridge School of Clinical Medicine, Cambridge, UK; Baillie Gifford Pandemic Science Hub, Centre for Inflammation Research, The Queen’s Medical Research Institute, The University of Edinburgh, Edinburgh, UK; Roslin Institute, The University of Edinburgh, Easter Bush, Edinburgh, UK; MRC Human Genetics Unit, Institute of Genetics and Cancer, The University of Edinburgh, Western General Hospital, Edinburgh, UK; Intensive Care Unit, Royal Infirmary of Edinburgh, Edinburgh, UK; UK Dementia Research Institute at the University of Edinburgh, Edinburgh, UK; Institute for Neuroscience and Cardiovascular Research, The University of Edinburgh, Edinburgh, UK

## Abstract

Cellular mechanisms in early Alzheimer’s disease (AD) remain poorly understood despite generation of extensive observational multiomic data. Weighted integration across diverse datasets can prioritise convergent mechanisms and targets. We conducted a systematic review to identify datasets profiling cell-type-specific changes in early AD. We integrated multiomic findings using Meta-Analysis by Information Content (MAIC), a computational method which combines genomic data from diverse experimental sources. Electron transport chain dysfunction in mitochondria emerged as a shared pathway in neurons, microglia, and astrocytes, and Wnt signalling was a prominent astrocyte-specific signature. Pathological neuro-glial signalling mechanisms included APP-CD74, APOE-SORL1, and WNT-FZD/LRP6. Integration with pharmacological data identified therapeutics already under investigation in AD, such as guanfacine, and candidates not previously prioritised, such as eltrombopag and encorafenib. We present a systematic analysis of cell-type-specific changes in early AD to prioritise pathways for investigation and therapeutics for repurposing.

## 1. Introduction

Alzheimer’s disease (AD), the most common cause of dementia, is a leading and growing cause of global disability and death.^1^ Effective treatments have historically been lacking. Recently, new therapeutics targeting β-amyloid deposition have demonstrated a proof-of-concept for developing effective treatments guided by disease genetics and mechanisms.^2^ However, clinical benefits from these medications remain small, and significant adverse events mean they are not yet widely implemented in routine clinical practice. Therefore, new, more effective treatments are urgently required.

AD is a progressive disease. There is emerging interest in understanding early disease mechanisms that occur prior to neurodegeneration and synapse loss, as interventions at this stage are most likely to be effective. The early cellular phase of AD is complex. In addition to neuronal dysfunction, non-neuronal cell-types, particularly microglia and astrocytes, undergo neuroprotective and deleterious changes linked to neurodegeneration. Given this complexity, it is likely that a multifaceted therapeutic strategy targeting diverse disease mechanisms across different cell-types will provide the most clinical benefit.^3^

The focus on delineating early cellular changes in AD is reflected by the rapid growth of system-wide data generated by high-throughput assays.^4^ These include transcriptomic and proteomic data generated from rodent models, as well as human studies, including genome-wide association studies (GWASs) and genomic analyses of post-mortem or surgically-resected AD brain tissue. These studies ultimately generate information in a consistent format: lists of genes or proteins implicated in a biological process. A significant challenge has become the integration of these diverse biological datasets to identify and prioritise mechanisms for therapeutic targeting.

In this study, we addressed this challenge by first conducting a systematic review to identify all relevant human and rodent transcriptomic and proteomic datasets profiling cell-type-specific changes in early AD. We then integrated these datasets, together with unbiased data from an AD GWAS and curated data from KEGG pathways, using Meta-Analysis by Information Content (MAIC).^5^ MAIC is a computational tool which we previously developed to specifically combine genomic data from diverse experiments without prior assumptions about the quality of individual data sources, and which we have shown to predict novel experimental findings.^6,7^ Using MAIC, we identified patient-relevant genomic changes in early AD which were conserved across experimental approaches, and present integrated changes in cell-type-specific gene expression, functional pathways, and intercellular signalling that prioritise targets for future therapeutic investigation.

## 2. Results

### 2.1. Systematic review identifies cell-type-specific early AD datasets

First, we conducted a systematic review to identify existing datasets examining cell-type-specific changes in early AD **(Figure 1).** The combined search for cell-type-specific datasets from early AD tissue identified 4,569 studies. We extracted 50 studies for full-text evaluation, of which 18 met the pre-defined criteria of describing cell-type-specific genomic data in early AD. Four additional studies were included from reference literature. In total, we identified 13 eligible datasets from neurons, 18 from microglia, and 12 from astrocytes **(Table 1, Supplementary Data)**. We also identified seven datasets from oligodendrocytes and four from brain endothelial cells, but these were insufficient for meta-analysis. Since MAIC learns from all input data, we decided *a priori* to seed the algorithm with high-quality data from the most recent meta-analysis of AD GWASs and the human AD KEGG list. This resulted in 45 unique gene lists being included across three analyses.

**Figure 1.**
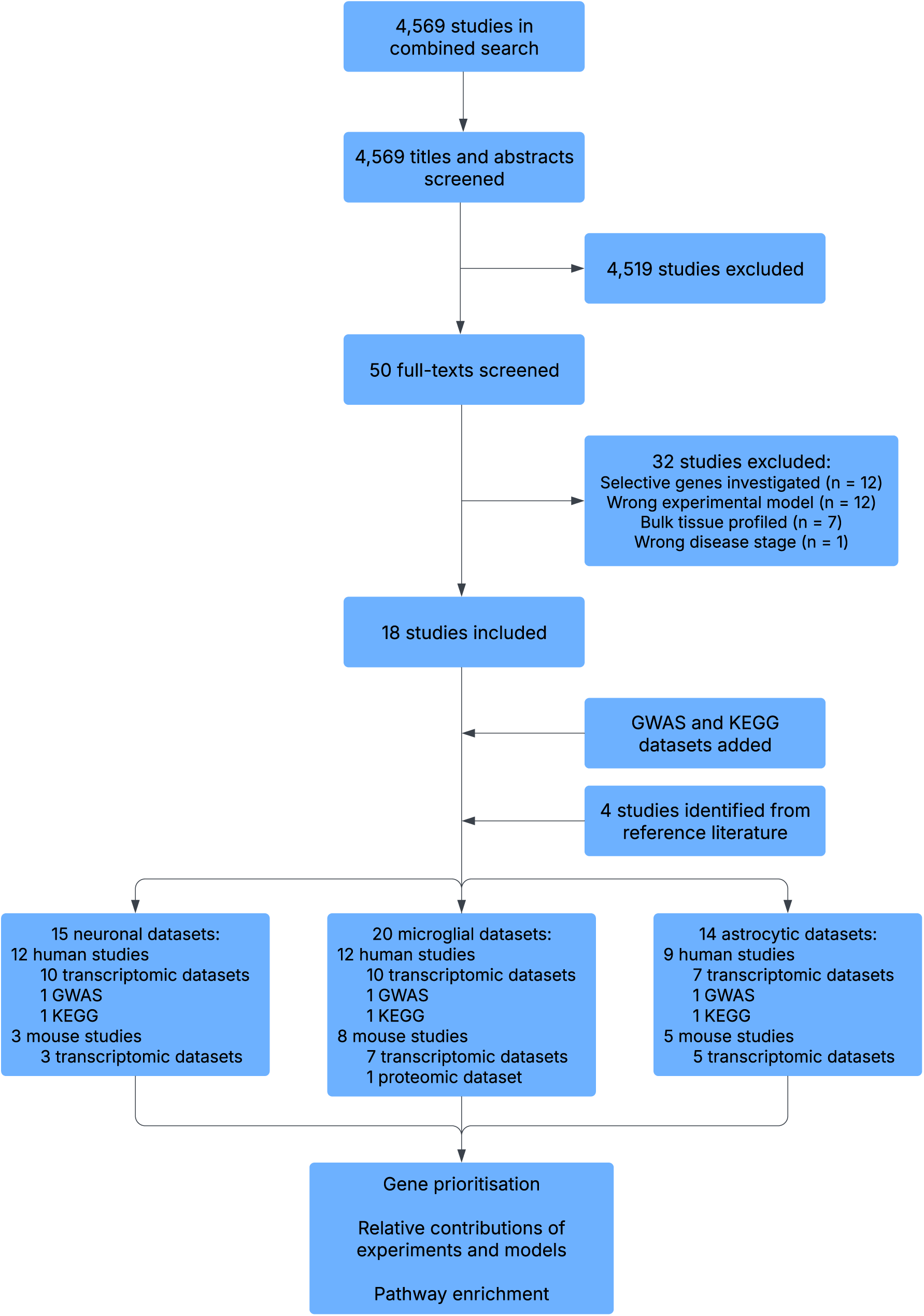
CONSORT diagram of included datasets. We included any human or rodent study describing differentially expressed genes or proteins in neurons, microglia or astrocytes from early-stage AD brain tissue compared to control. GWAS = genome-wide association study, KEGG = Kyoto Encyclopaedia of Genes and Genomes.

**Table 1.** Description of the datasets included in MAIC analyses. AD = Alzheimer’s disease, WT = wild-type, DEGs = differentially expressed genes, TRAP = translating-ribosome-affinity purification, DEPs = differentially expressed proteins, KEGG = Kyoto Encyclopaedia of Genes and Genomes.

| **Citation** | **Description** | **Cell-type isolation technique** | **Analyses** |
| --- | --- | --- | --- |
| Bellenguez Nat Genet 2022<sup>75</sup> | GWAS meta-analysis of AD | - | Neurons, microglia, astrocytes |
| Chen Cell 2020<sup>82</sup> | Spatial transcriptomics of various cell-types in AD vs WT mice at three months | RNAscope puncta using marker genes | Neurons, microglia, astrocytes |
| Depp Nature 2023<sup>83</sup> | DEGs in microglia in AD vs WT mice at six months | Magnetic-activated cell sorting | Microglia |
| Gazestani Cell 2023<sup>84</sup> | DEGs in various cell-types in patients with idiopathic normal pressure hydrocephalus and β-amyloid pathology vs those without β-amyloid pathology | 10X Genomics | Neurons, microglia, astrocytes |
| Gerrits Acta Neuropath 2021<sup>9</sup> | DEGs in microglia in an early cluster in patients with AD | Fluorescence-activated cell sorting | Microglia |
| Habib Nat Neuro 2020<sup>85</sup> | DEGs between disease-associated and homeostatic astrocytes in AD vs WT mice at seven months | 10X Genomics | Astrocytes |
| Jiwaji Nat Comms 2022<sup>86</sup> | DEGs in astrocytes in AD vs WT mice at six months using TRAP-seq | Crossing AD and Aldh1l1_eGFP-RPL10a mice, which specifically express GFP-tagged ribosomes in astrocytes | Astrocytes |
| KEGG_HSA05010<sup>8</sup> | KEGG list for human AD | - | Neurons, microglia, astrocytes |
| Keren-Shaul Cell 2017<sup>38</sup> | DEGs between disease-associated and homeostatic microglia in AD vs WT mice at three months | Fluorescence-activated cell sorting | Microglia |
| Kim Mol Neurodegen 2022<sup>87</sup> | DEGs in microglia in AD vs WT mice at six months | 10X Genomics | Microglia |
| Leng Nat Neuro 2021<sup>88</sup> | DEGs in various cell-types in patients with early AD vs control | 10X Genomics | Neurons, microglia, astrocytes |
| Li Glia 2020<sup>89</sup> | DEGs in astrocytes in AD vs WT mice at three months | Fluorescence-activated cell sorting | Astrocytes |
| Mancuso Nat Neuro 2024<sup>90</sup> | DEGs in xenotransplanted human stem-cell-derived microglia in AD vs WT mice at six months | Fluorescence-activated cell sorting | Microglia |
| Mathys Cell 2023<sup>91</sup> | DEGs in various cell-types in patients with early global AD pathology | 10X Genomics | Neurons, microglia, astrocytes |
| Mathys Cell Rep 2017<sup>92</sup> | DEGs between early response and homeostatic microglia in CK-p25 vs control mice at one week | Fluorescence-activated cell sorting | Microglia |
| Mathys Nature 2019<sup>39</sup> | DEGs in various cell-types in patients with early AD pathology vs control | 10X Genomics | Neurons, microglia, astrocytes |
| Monasor eLife 2020<sup>93</sup> | DEPs in microglia in AD vs WT mice at three months | Magnetic-activated cell sorting | Microglia |
| Olah Nat Comms 2020<sup>94</sup> | DEGs upregulated in early response microglia in patients with AD vs control | Fluorescence-activated cell sorting | Microglia |
| Serrano-Pozo Nat Neuro 2024<sup>95</sup> | DEGs in astrocytes clustering in early gene sets in patients with AD | 10X Genomics | Astrocytes |
| Simpson Neurobio Ag 2011<sup>10</sup> | DEGs in astrocytes in patients with moderate vs low AD pathology | Laser-capture microdissection | Astrocytes |
| Sobue Acta Neuro Comms 2021<sup>96</sup> | DEGs in microglia in AD vs WT mice at eight months | Magnetic-activated cell sorting | Microglia |
| Sun Cell 2023<sup>97</sup> | DEGs in early response microglia in patients with AD vs control | 10X Genomics | Microglia |
| Wachter Acta Neuro 2024<sup>98</sup> | DEGs in microglia in patients with medium vs low β-amyloid pathology | 10X Genomics | Microglia |
| Zhou Nat Med 2020<sup>99</sup> | DEGs in various cell-types in patients with AD and in AD vs WT mice at seven months | 10X Genomics | Neurons, microglia, astrocytes |

There were 29 (64%) human and 16 (36%) mouse datasets. Most gene lists came from transcriptomic experiments (n = 42, 93%) and there was one proteomic dataset (2%). Two (4%) gene lists (KEGG and one human microglial transcriptomic dataset [Gerrits et al. 2021]) were unranked.^8,9^

The earliest included study was published in 2011 (Simpson et al. 2011), but 83% (n = 19/23) of studies were published since 2020.^10^ We performed MAIC analyses using these datasets and used outputs from MAIC to prioritise genes, understand the relative contributions of different experimental approaches, and identify enriched pathways and cell-cell communication mechanisms.

### 2.2. MAIC prioritises cell-type-specific changes in early AD across integrated datasets

We used MAIC to identify cell-type-specific changes in neurons, microglia, and astrocytes consistently implicated across diverse datasets and to prioritise highly-ranked genes (**Figure 2a**). Most highly-ranked genes were supported by human and mouse transcriptomic lists due to the abundance of these sources but were also identified in GWAS and/or KEGG data, leading to their prioritisation in the MAIC algorithm.

**Figure 2.**
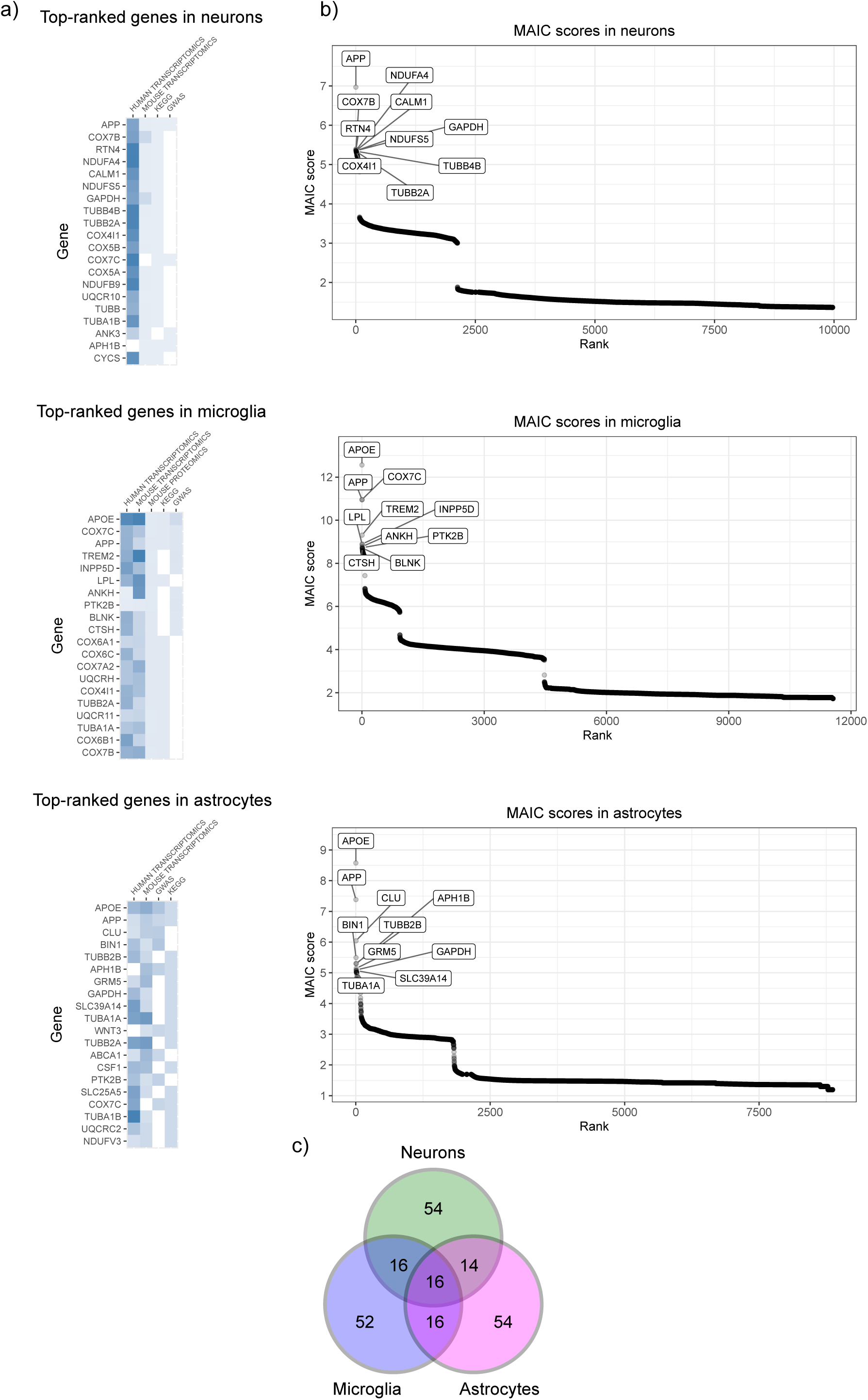
Highest ranked genes in each cell-type and distribution of MAIC scores by gene rank. a) Heatmaps display the 20 top-ranked genes in each cell-type. For each gene, the summed MAIC scores from each data source are highlighted. b) MAIC scores across ranked genes. Genes which rank highly in lists from different data sources have the highest MAIC scores. Genes with baseline MAIC scores are supported by limited evidence. c) Overlap between the top 100 genes of each cell-type-specific MAIC analysis. KEGG = Kyoto Encyclopaedia of Genes and Genomes, GWAS = genome-wide association study.

In neurons, APP, COX7B, RTN4, NDUFA4, and CALM1 were the five highest-ranked genes (**Figure 2b**) and were consistently supported by strongly weighted transcriptomic studies in humans, but only weakly by the unbiased data from a large-scale GWAS. In microglia and astrocytes, a striking species difference was evident. Many highly-ranked genes in microglia, including APOE, TREM2, and LPL, were disproportionately supported by mouse transcriptomic data, despite this being a less commonly included experimental category. In astrocytes, APP, CLU, APH1B, and GRM5 were disproportionately represented in transcriptomic data from mice.

An analysis of the top 100 genes identified in each cell-type-specific MAIC analysis found that 16 genes were shared between all three cell-types and a further 14–16 genes overlapped between two cell-types (**Figure 2c**). Therefore, >50% of the top 100 genes were unique to each cell-type, demonstrating the cell-type-specificity of pathological changes in early AD.

### 2.3. Human transcriptomic data contributes most to MAIC scores across cell-types

The relative contribution of different data sources to MAIC scores reflects both the number of studies in each category and the amount of overlap (corroboration) between one category and the others (**Figure 3**). Human transcriptomic data made the largest contribution to the summed MAIC scores of the top 20 genes in each cell-type, but this was particularly significant in neurons (72%), while microglia (38%) and astrocytes (38%) had more balanced sources of evidence. This is partly explained by the proportion of gene lists in the neuronal analysis coming from human transcriptomic experiments. These findings were also reflected by analyses of the contributions of individual studies to overall MAIC scores in each cell-type (**Supplementary Fig. 1**).

**Figure 3.**
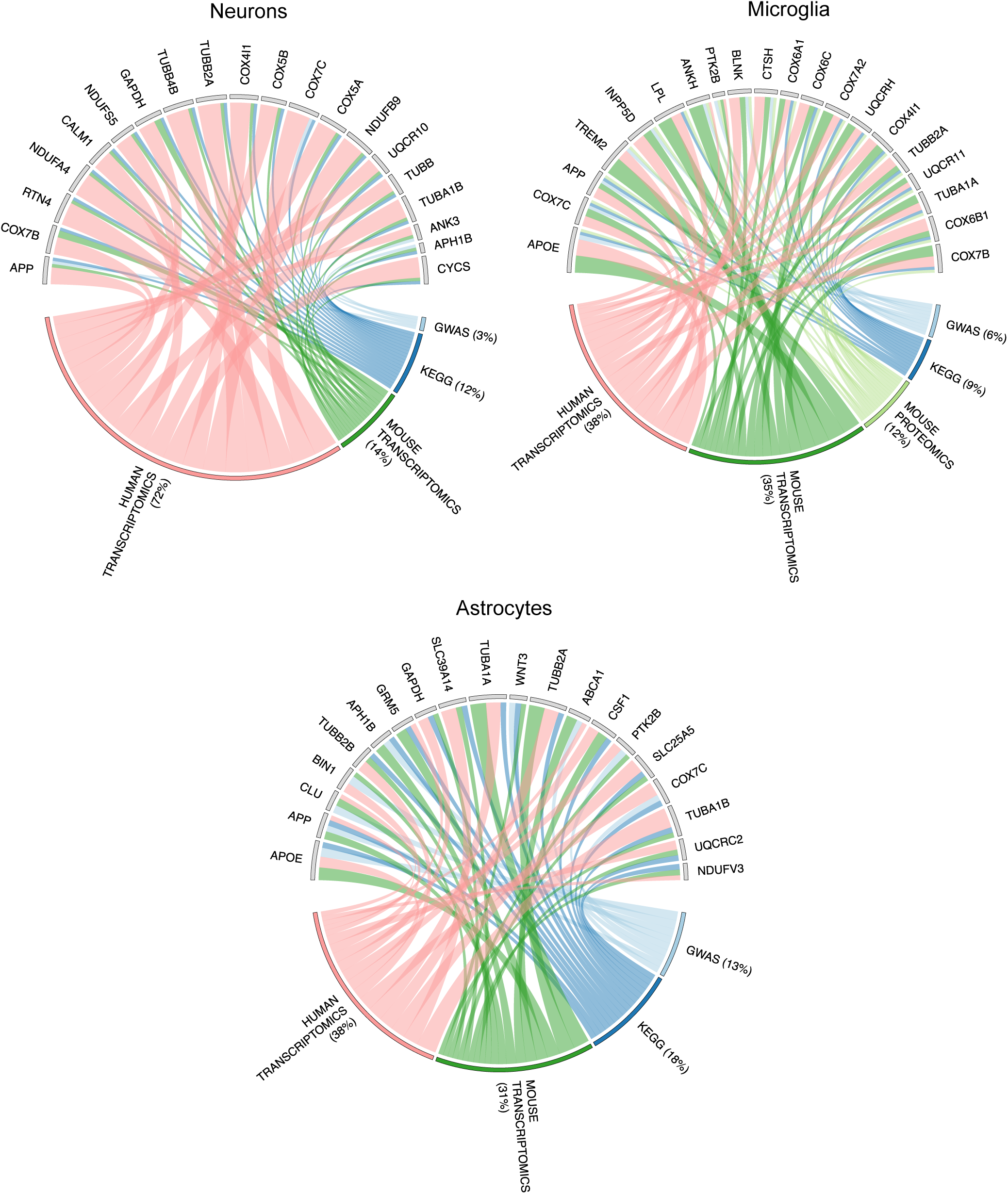
Relative contribution of different data sources to the top 20 genes in each cell-type. The size of blocks for each data source and gene is proportional to their summed MAIC scores. Percentages represent the contribution of each data source to the total MAIC scores of the top 20 genes. For each gene, connecting lines show the data source origins of its summed MAIC score. KEGG = Kyoto Encyclopaedia of Genes and Genomes, GWAS = genomewide association study.

Since GWAS data is less susceptible to biases introduced by assays and experimental conditions, and reflects the underlying causes of disease susceptibility, we further analysed the contribution of GWAS data to the top 20 genes in each cell-type. GWAS data made a greater contribution to summed MAIC scores in the top 20 genes in astrocytes (13%) and microglia (6%) than in neurons (3%; **Figure 3**). In addition, nine in each of the top 20 genes in microglia (APOE, COX7C, APP, TREM2, INPP5D, ANKH, PTK2B, BLNK, and CTSH) and astrocytes (APOE, APP, CLU, BIN1,

APH1B, WNT3, ABCA1, PTK2B, and COX7C) were supported by the GWAS, compared to four in the top 20 genes in neurons (APP, COX7C, ANK3, and APH1B).

### 2.4. Protein-protein and ligand-receptor interaction analyses identify cell-autonomous and non-cell-autonomous signalling

Alterations in protein-protein interactions and cell-cell crosstalk have been proposed to drive both cell-autonomous and non-cell-autonomous pathology in AD. To understand changes in intra- and intercellular signalling, we identified protein-protein interaction networks and ligand-receptor pairs between the top 100 genes ranked by MAIC score in each cell-type (**Figure 4a**). Across all cell-types, we identified a large cluster of genes involved in aerobic electron transport and mitochondrial ATP synthesis. In neurons, other highly-ranked clusters were related to APP metabolism and microtubule trafficking. In microglia, there were prominent immune signatures, including viral mRNA translation and B cell receptor activation. In astrocytes, we identified groups of genes involved in lipoprotein binding, AMPK activation, and Wnt signalling. Rank-based KEGG and Reactome analyses described similar shared and cell-type-specific signatures (**Supplementary Fig. 2**).

**Figure 4.**
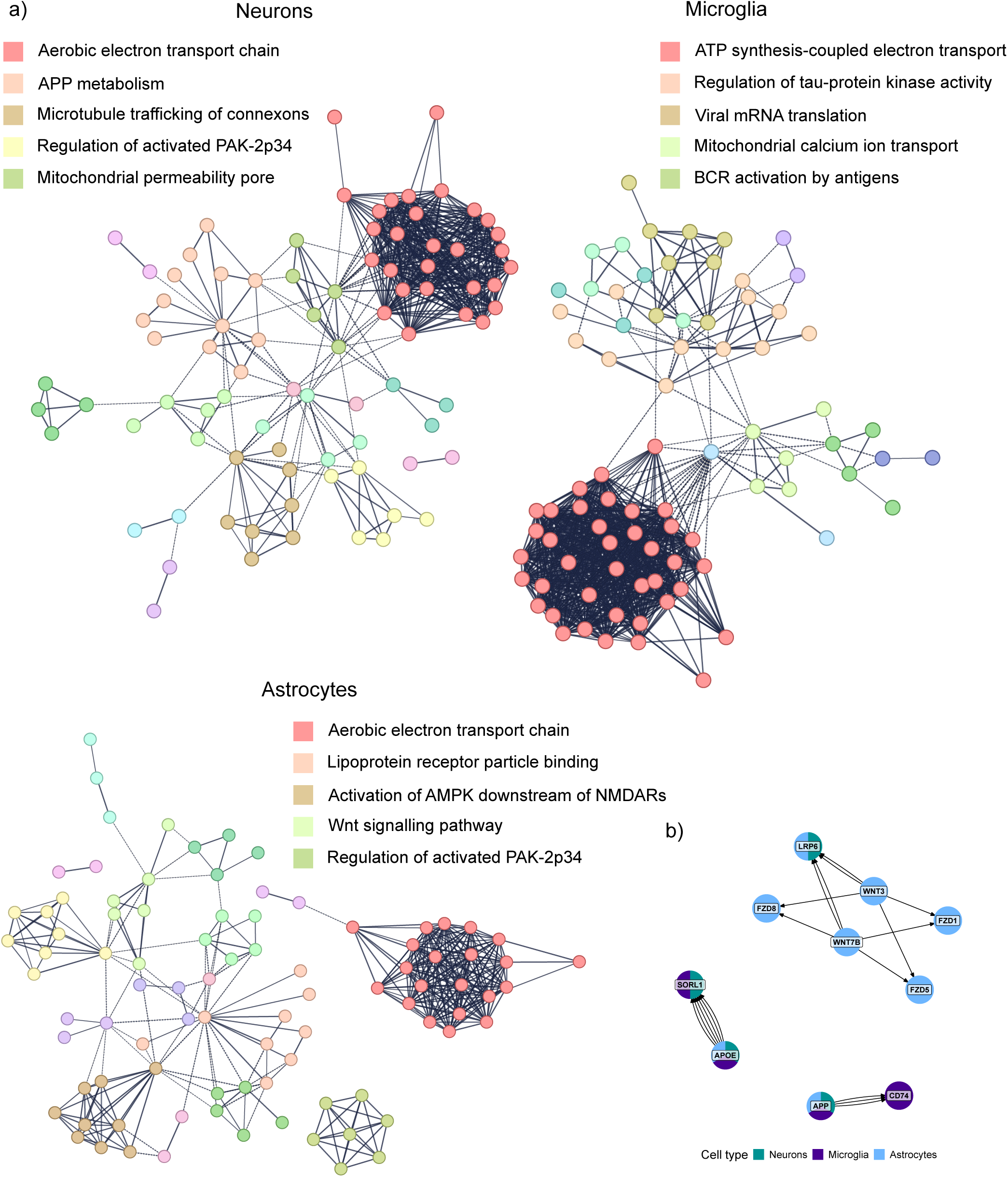
Intra- and intercellular communication between the top 100 genes in each cell-type. a) The five largest protein-protein interaction networks are labelled in each analysis. Solid lines represent interactions within networks, and their thickness is proportional to the confidence in their interaction. Dotted lines represent interactions between networks. b) Ligands point toward their corresponding receptors and nodes are coloured according to the cell-type analyses in which they appeared.

Next, we searched an open-source ligand-receptor database to identify potential cellular crosstalk mechanisms between the top 100 genes in each cell-type (**Figure 4b**). APP-CD74 was a ligand-receptor pair between all three cell-types and microglia and APOE-SORL1 was a pair between all three cell-types and neurons and microglia. There were several Wnt signalling pairs among astrocytes and between astrocytes and neurons, including WNT3/7B-LRP6 (astrocytes-neurons and astrocytes-astrocytes) and WNT3/7B-FZD1/5/8 (astrocytes-astrocytes).

### 2.5. Drugs and compounds targeting proteins encoded by the top 100 genes in each cell-type

Finally, we aimed to identify therapeutics whose targets were enriched among the top 100 genes in each cell-type (**Figure 5**). We identified 22 significantly enriched (adjusted p-value < 0.05) drugs or compounds in neurons, 47 in microglia, and 30 in astrocytes. Our prioritised list included drugs already under investigation in pre-clinical or clinical AD contexts, supporting the use of our methodology to validate existing therapeutics (**Figure 5a**). Clinical-stage drugs included guanfacine (adjusted p-value = 5.90 × 10^-9^, odds ratio = 20.9 in neurons; adjusted p-value = 2.79 × 10^-10^, odds ratio = 23.1 in microglia), desipramine (adjusted p-value = 5.63 × 10^-8^, odds ratio = 18.9 in neurons; adjusted p-value = 2.91 × 10^-9^, odds ratio = 21.1 in microglia), and minocycline (adjusted p-value = 9.87 × 10^-7^, odds ratio = 16.3 in neurons; adjusted p-value = 1.70 × 10^-5^, odds ratio = 14.4 in microglia). Pre-clinical compounds included MRS 2578, a P2Y_6_R antagonist (adjusted p-value = 0.01, odds ratio = 10.7 in microglia); AMN 082, a mGluR7 agonist (adjusted p-value = 0.01, odds ratio = 10.7 in microglia; adjusted p-value = 4.95 × 10^-3^, odds ratio = 10.8 in astrocytes); CA-074, a cathepsin B inhibitor (adjusted p-value = 0.01, odds ratio = 9.01 in astrocytes); and STF-31, a GLUT1 inhibitor (adjusted p-value = 0.04, odds ratio = 7.65 in neurons).

**Figure 5.**
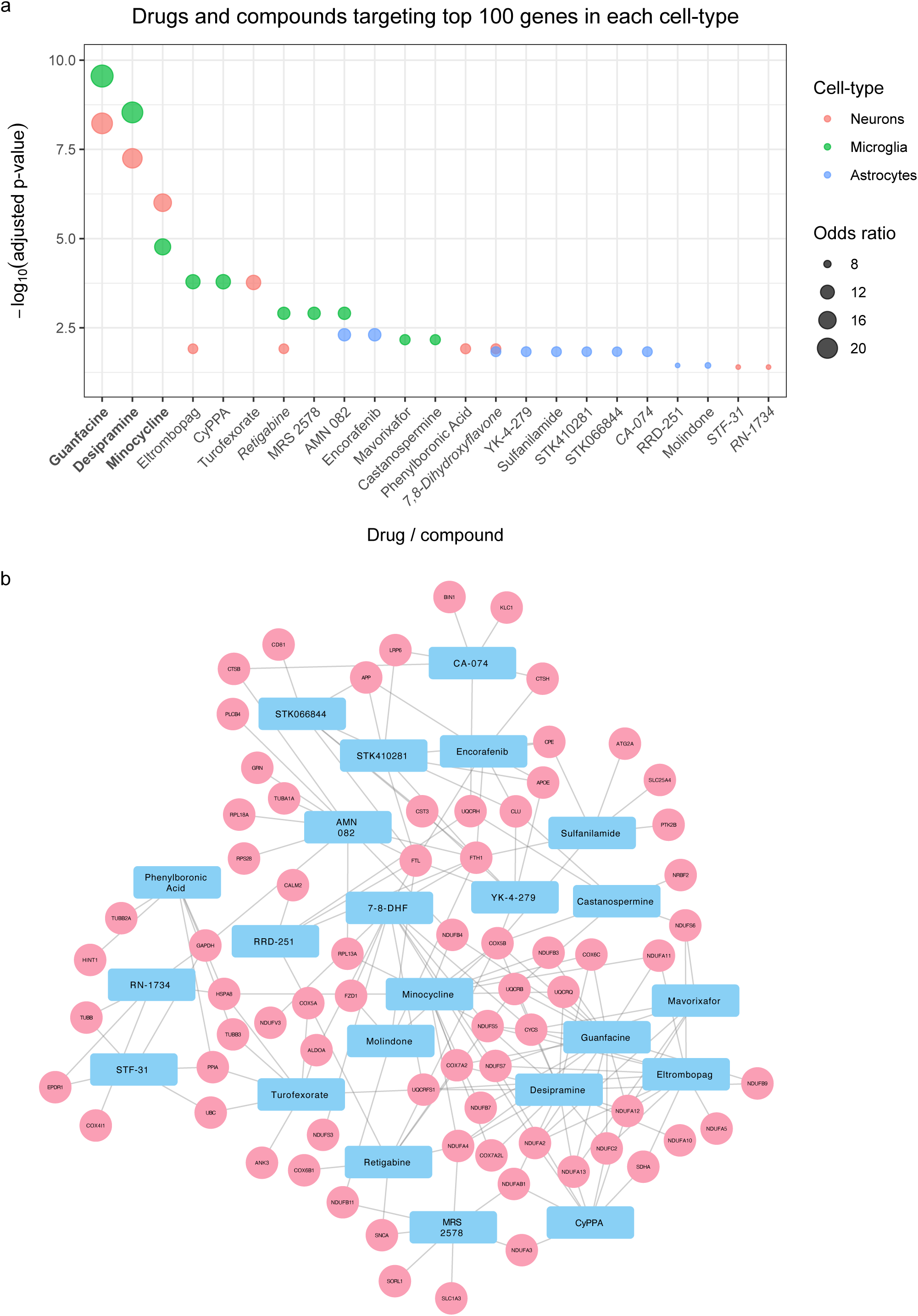
Drugs and compounds targeting proteins encoded by the top 100 genes in each cell-type. a) Top 10 drugs and compounds enriched for target proteins encoded by the top 100 genes in each cell-type. Drugs in bold have been studied in patients with Alzheimer’s disease and drugs in italics have been studied in pre-clinical models of Alzheimer’s disease. b) Network diagram displaying the top-ranked genes in each cell-type which are targeted by prioritised drugs and compounds.

Importantly, our prioritised list also included licensed drugs which have not previously been studied in the context of AD, such as eltrombopag, a thrombopoietin receptor agonist used to treat thrombocytopaenia and aplastic anaemia (adjusted p-value = 0.01, odds ratio = 8.94 in neurons; adjusted p-value = 1.62 × 10^-4^, odds ratio = 12.4 in microglia); and encorafenib, a BRAF kinase inhibitor used to treat melanoma (adjusted p-value = 4.95 × 10^-3^, odds ratio = 10.8 in astrocytes) (**Figure 5a**).

## 3. Discussion

We present a comprehensive, data-driven meta-analysis to identify and prioritise cell-type-specific changes in early AD which are consistent across diverse experimental approaches. We used a systematic approach to identify datasets, selecting those which were cell-type-specific to minimise the artefacts and loss-of-signal seen in bulk-tissue analyses. This strategy increases our confidence that these mechanisms reflect biologically-relevant processes in early AD and allows us to prioritise existing drugs and molecules for further investigation.

Our meta-analysis highlights AD-associated changes in mitochondrial function, particularly involving electron transport chain, as a top-ranked cluster across all three cell-types. The causal role of mitochondrial dysfunction in AD is supported by a wide range of pre-clinical and clinical studies. Variants in COX6B1, NDUFA4, SURF1, and COX10 have been associated with AD and expression of these genes correlates with β-amyloid plaque burden in the mouse hippocampus.^11^ A multiomic study of the human hippocampus demonstrated that electron transport chain gene expression decreases in astrocytes during ageing.^12^ In microglia in ageing mice, mitochondrial DNA leakage into the cytosol activates the cGAS-STING pathway and prostaglandin E2 signalling limits mitochondrial respiration, both of which drive inflammation and neurodegeneration.^13–15^ Genetic and pharmacological inhibition of mitochondrial function, including of the electron transport chain complexes I, III and IV, increases APOE expression.^16,17^ Notably, reduced expression of respiratory complex I and COX7C preceded observed increases in APOE expression in a mouse model of AD. COX7C ranked within the top 20 genes in all three of our analyses and SLC25A genes, which encode mitochondrial membrane transporters, ranked within the top 35 genes in all three cell-types. Conversely, promoting mitochondrial function using a designer receptor exclusively activated by designer drugs (DREADD) in a mouse model of AD rescued cognitive impairment.^18^ Mechanistically, across pre-clinical models, tau enters mitochondria in neurons and interacts with NDUFS3, which is part of complex I and ranked 38 in our neuronal analysis, leading to reverse electron transport. This produces reactive oxygen species, which perpetuate tau hyperphosphorylation in a feedforward loop, and inhibiting this mechanism rescued neurodegeneration across models of tauopathy.^19^ Reactive oxygen species in neurons also lead to the accumulation of lipid droplets in glia during early neurodegeneration.^20^ In the clinical setting, functional imaging using positron-emission tomography confirmed that patients with early AD have decreased mitochondrial complex I expression which progresses in the early disease course and correlates with reduced cognitive performance.^21^ Together, these results suggest that mitochondrial dysfunction, particularly via altered electron transport chain activity, plays an important role in the early stages of AD and are consistent with this being a top-ranked pathway in our neuronal and glial analyses.

In astrocytes, we identified an enrichment of genes involved in Wnt signalling, including ligand-receptor pairs such as WNT3/7B-LRP6 and WNT3/7B-FZD1/5/8 among highly-ranked genes. The Wnt signalling pathway is required for synaptic function, including synaptogenesis in postnatal development, and deficient Wnt signalling is associated with synapse loss in AD.^22,23^ During development, microglia-mediated synaptic engulfment, which also occurs in early AD, depends on Wnt signalling to astrocytes.^24,25^ Dickkopf-1 (DKK1) is a Wnt antagonist that is required for β-amyloid-induced synapse loss in AD and blocking its action is protective.^26–28^ In addition, three variants in LRP6 have been associated with late-onset AD.^29,30^ An isoleucine to valine substitution in LRP6 (LRP6-Val) is a common variant in European populations and LRP6-Val knock-in mice display defective Wnt signalling and synaptic impairment.^31,32^ Mechanistically, this pathology is caused by an altered Wnt7a-mediated interaction between LRP6 and FZD5. Further supporting a role for aberrant Wnt signalling in AD, a proteomic study of CSF from patients with AD identified early changes in the expression of soluble β-catenin (CTNNB1) and AXIN2, a suppressor of Wnt/ β-catenin signalling which affects mitochondrial biogenesis.^33^ Phosphorylated β-catenin has been shown to accumulate in the hippocampal pyramidal neurons of patients with AD, a signature that was replicated with inhibition of the proteasome.^34^ Our data extend these findings to support a role for dysfunctional Wnt signalling, including via LRP6, in the early stages of AD and suggest astrocytes may play a key role in this pathological mechanism.

Our integrated analyses also identify several potentially important ligand-receptor interactions, prioritising targets for therapeutic intervention. APP-CD74 was a pair between the top 100 genes of all three cell-types and microglia. CD74 encodes a membrane protein that chaperones MHCII molecules and acts as a receptor for macrophage migration inhibitory factor (MIF).^35^ Its expression is increased in microglia, astrocytes, tangle-positive neurons, and β-amyloid plaques in human AD tissue.^36^ Microglia which express high levels of CD74 in AD adopt morphological characteristics associated with microglial activation.^37^ In single-cell RNA sequencing datasets, increased CD74 expression in microglia is associated with reduced expression of homeostatic markers and is positively correlated with expression of “activated response” genes following exposure to β-amyloid plaques.^38,39^ Increased CD74 expression in microglia has also been identified in brain tissue from patients with multiple sclerosis.^40^ *In vitro*, CD74 inhibits β-amyloid production, potentially by regulating APP trafficking in neurons.^41^ In addition, lipopolysaccharide (LPS) challenges *in vitro* and in mice increase microglial expression of CD74.^42^ In a mouse model of AD, treatment with a molecule which binds CD74 to inhibit MIF improved cognition and mitochondrial function, though without altering β-amyloid plaque burden.^43^

Similarly, APOE-SORL1 was a ligand-receptor pair between the top 100 genes of all three cell-types and neurons and microglia. A recent transcriptomic study found APOE-SORL1 and APOE-TREM2 to be top-ranked signalling axes in human AD microglia and associated with disease progression.^44^ SORL1 has been identified in late-onset AD GWASs and rare variants in SORL1 are also known to cause early-onset AD.^45,46^ SORL1 is involved in intracellular trafficking, including of APP, which accumulates in endosomes when expression of SORL1 is lost.^47,48^ In our analyses, intracellular trafficking was a highly-ranked protein-protein interaction network in neurons. In hiPSC-derived neurons, SORL1 deletion has been shown to reduce APOE and CLU expression, while in astrocytes, SORL1 knockout increases the expression of both of these AD-associated genes.^49^ In neurons, this leads to an increase in β-amyloid and phosphorylated tau levels. SORL1 and APOE mRNA and protein levels also correlate in neurons from post-mortem human brain tissue. In iPSC-derived astrocytes, SORL1 co-precipitates with APOE, and SORL1 knockout in these cells is also associated with accumulation of β-amyloid.^50^ Our findings highlight the possibility of neuro-glial crosstalk via APOE-SORL1 as a pathological mechanism in the early stages of AD.

A key strength of our work is that many of the top-ranked genes identified using MAIC derive predominantly from human datasets, enhancing the translational potential of our findings. In addition, overlap between MAIC-prioritised genes and population-level studies further substantiates the clinical relevance of our findings. This includes a recent Mendelian randomisation analysis which used the UK Biobank and deCODE Health Study to identify causal associations between plasma protein concentrations and neurodegenerative disorders, including AD.^51^ The study found 20 unique proteins to be associated with AD and supported by co-localisation, including eight previously unreported loci. Of these 20 proteins, three (APOE, BLNK, and CTSH) ranked within the top 10 genes of our microglial analysis and a further two (GRN and CD2AP) ranked within the top 100 genes. Two proteins (APOE and CTSH) ranked within the top 30 genes of our astrocytic analysis and two (MME and APOE) in the top 80 of our neuronal analysis. The overrepresentation of genome-wide AD associations in glial cell-types is supported by our analysis of experimental contributions to MAIC scores and has previously been shown in cell-type enrichment analyses.^52^ CTSH is a member of a group of proteases located in lysosomes that has been reported in AD GWASs.^53^ CTSH expression is increased in the brain tissue of patients with AD and its knockout increases microglial phagocytosis of β-amyloid peptides. This suggests that protective variants in CTSH are associated with reduced gene expression and facilitate microglial clearance of β-amyloid, a key pathological mechanism in early AD. CD2AP and MME are likewise supported by AD GWAS data and are involved in the clearance of β-amyloid.^54,55^ Our results therefore provide a method of validating findings from genome-wide studies to support the involvement of specific genes in early AD at cell-type resolution.

Finally, we describe existing drugs and compounds whose targets were enriched among highly-ranked genes in each cell-type. Recent work validated this approach of using single-cell transcriptomic data to identify existing therapeutics for repurposing in AD.^56^ Our findings include therapies that are currently in clinical trials for patients with AD, such as guanfacine, which was the top-ranked drug in both our neuronal and microglial analyses.^57^ Guanfacine is an α-2A adrenergic receptor agonist approved for treating attention deficit hyperactivity disorder (ADHD) and has been proposed to target noradrenergic dysfunction, which occurs early in AD due to pathology affecting the locus coeruleus.^58^ Minocycline, which ranked third in both neurons and microglia, is a tetracycline antibiotic with anti-inflammatory properties. It has produced promising pre-clinical results and was recently tested in a phase II clinical trial in mild AD.^59^

We identified several compounds which target key neuro-glial mechanisms in AD and are supported by pre-clinical evidence. MRS 2578 antagonises the G-protein coupled receptor P2Y_6_R, which is activated by stressed but viable neurons and mediates microglial phagocytosis of neurons in response to β-amyloid and tau.^60–62^ Pharmacological inhibition and genetic knockdown of P2Y_6_R have been shown to prevent loss of viable neurons and improve cognition in models of AD. AMN 082 is a mGluR7 agonist which inhibits the release of glutamate and protects against NMDA receptor-mediated excitotoxicity in models of neuronal death.^63–65^ β-amyloid impairs these protective functions of mGluR7 signalling in the basal forebrain, a region of early degeneration in AD.^66^ CA-074 inhibits cathepsin B, a cysteine protease which facilitates β-amyloid degradation in lysosomes and microglia but can also adopt β-secretase-like activity to promote the production of β-amyloid plaques.^67,68^ In AD, lysosomal membrane permeabilisation can cause cathepsin B to localise within the cytosol, where it activates the NLRP3 inflammasome, leading to the release of IL-1-β and IL-18.^69,70^ In AD models with wild-type β-secretase sites, CA-074 improves memory deficits.^71,72^ Finally, STF-31 inhibits GLUT1, which is highly expressed by microglia in inflammatory conditions, and has been shown to reduce photoreceptor loss in a model of retinal degeneration.^73,74^

Importantly, our integrated meta-analysis also highlights candidate therapeutic targets not previously investigated in AD. Notably, we prioritise eltrombopag, a thrombopoietin receptor agonist licensed for thrombocytopaenia and aplastic anaemia, and encorafenib, a BRAF kinase inhibitor used for melanoma, for repurposing. Since both drugs are already in clinical use, they offer credible avenues for rapid translational evaluation. These results support the feasibility of integrating multimodal genomic evidence to identify existing therapeutics for repurposing to AD.

The primary strength of this work lies in its integration of diverse datasets. This mitigates biases inherent to individual model systems and enhances generalisability. MAIC has been specifically designed to integrate data from diverse genomic and proteomic sources and has been shown to be superior to alternative methods.^6^ MAIC further benefits from being able to incorporate both ranked and unranked data through unbiased, data-driven weightings.

Our work has several limitations. Our analysis was restricted to datasets presenting large gene sets rather than single-gene or candidate gene studies. Therefore, some relevant findings may have been missed. In addition, our analyses were confined to neurons, microglia, and astrocytes. Oligodendrocytes and other cell-types were excluded due to insufficient numbers of datasets and their addition in future may provide additional insights. Third, the rank-based nature of the meta-analysis did not account for the direction of changes in gene expression. Fourth, our analysis only included datasets identified in the systematic review conducted in December 2024. This is a rapidly evolving field and subsequently published studies have not been captured. Future work aims to develop a live systematic review and MAIC pipeline to enable prompt incorporation of newly available data. Finally, while our analyses of enriched protein-protein interaction networks and pathways provide some insight, functional characterisation of the contributions of highly-ranked genes to early AD pathology is required. Future studies should focus on identifying novel therapeutic targets for early AD using the highly-ranked genes in our analyses.

In conclusion, there has been a recent rapid expansion in cell-type-specific, multiomic datasets generated from humans and animal models with the early stages of AD. Here, we provide a data-driven meta-analysis of these studies to facilitate prioritisation of their findings in neurons, microglia, and astrocytes. We identified cross-validated changes in the mitochondrial electron transport chain across all three cell-types and in Wnt signalling in astrocytes. Among highly-ranked genes, we found several potentially important ligand-receptor pairs, including APP-CD74, APOE-SORL1, and those involved in Wnt signalling, which may serve as mechanisms for early pathological neuro-glial crosstalk in AD. Finally, using these gene sets, we prioritise existing drugs and compounds for further investigation. These findings provide insight into the cellular and molecular signatures of early AD and identify new putative drug targets.

## 4. Methods

### 4.1. Literature search and systematic review

Studies and datasets were identified on PubMed and GEO NCBI using the search strategy described below in December 2024. We included any human or rodent study describing differentially expressed genes or proteins in neurons, microglia or astrocytes from early-stage AD brain tissue compared to control. Early AD was defined by pathological criteria (e.g. Braak stage) or age in cases of specific animal models (e.g. < 9 months for APP/PS1 mice). To provide a degree of background information on genes known to be involved in the pathogenesis of AD, we also included a recent AD GWAS and an unranked list of genes included in the human AD Kyoto Encyclopaedia of Genes and Genomes (KEGG) list (HSA05010) in our MAIC analyses.^8,75^ We excluded studies of late AD, bulk brain tissue, and selected subsets of genes.

The search terms were: (“Alzheimer”) AND (neuron* OR microglia* OR astrocyt* OR oligo* OR endothel*) AND (early[Title/Abstract] OR preclinical[Title/Abstract]) AND ((gene*[Title/Abstract]) OR (genom*[Title/Abstract]) OR (transcript*[Title/Abstract]) OR (protein*[Title/Abstract]) OR (susceptib*[Title/Abstract])) NOT (Review[Publication Type]).

### 4.2. Meta-analysis

MAIC (baillielab.net/maic/) combines ranked and unranked gene lists from diverse experimental sources to prioritise genes which are consistently identified by a set of related experiments, without making prior assumptions about the quality of individual data sources.^5^ MAIC is an iterative optimisation function between two simple heuristics: 1) a gene is more likely to be truly involved in a biological process if it appears in multiple experiments; and 2) an experiment that prioritises well-corroborated genes is a more reliable source of information. MAIC calculates a weighting factor for each experiment, which is used to assign a score to each gene. These scores are used to produce a ranked list of all genes included in the analysis. The MAIC algorithm has been shown to consistently outperform other methods for data integration of real biological data.^5^

We completed three MAIC analyses (neurons, microglia, and astrocytes). Datasets from different neuronal subtypes (e.g. excitatory neurons, inhibitory neurons, and interneurons) were combined in the neuronal analysis. Genes were ranked by their associated p-value (where available) or fold change. All genes (up to a pre-defined cut-off of 2000 genes) from ranked lists were included in the MAIC analyses (i.e. p-value or fold change cut-offs were not used). This was to ensure that genes approaching significance in multiple studies were included in our analyses. Where differential expression analyses between early AD and control were not publicly available, we analysed processed sequencing data using edgeR.^76^ Mouse genes were mapped to their human homologues using gProfiler, or excluded from the analysis if no homologue could be identified.^77^ Repeat occurrences of the same gene in a single gene list were removed. MAIC allows users to manually classify gene lists, for example by their experimental category. We categorised datasets as either human or mouse and then further defined the experimental methods as transcriptomics, proteomics, GWAS or KEGG.

### 4.3. Interpretation of MAIC outputs

First, we studied the distribution of MAIC scores in each of the three cell-type-specific analyses. Next, we analysed the relative contribution of different experimental approaches to the MAIC scores of individual genes and the overall ranked gene lists. To quantify relative contribution, we summed the MAIC scores of each gene from each experimental category and compared the contribution of each category to the MAIC scores of the top 20 genes. We further studied the information content of individual studies by summing the MAIC scores assigned to each of their genes and then normalising by the number of genes in the included list. We used STRING, which identifies protein-protein interactions, and Talkien, which identifies ligand-receptor pairs between gene lists, to perform interaction analyses within and between the top 100 genes in each analysis.^78,79^ To identify protein-protein interaction clusters using STRING, we used the Markov Clustering (MCL) algorithm with a minimum interaction score of 0.7 (high) and an inflation parameter of 3. Both tools extract curated and experimental data from a range of protein-protein interaction and cell-cell communication databases. We used the Wilcoxon rank-sum test to identify enriched KEGG and Reactome pathways among genes ranked by their MAIC score in each cell-type. Finally, we used Enrichr and the Proteomics Drug Atlas (2023) to identify existing drugs and compounds whose targets were enriched among the top 100 genes in each cell-type.^80,81^ R version 4.3.3 was used for data analysis and visualisation.

## Author contributions

MS: conceptualisation, methodology, formal analysis, investigation, data curation, writing – original draft, visualisation

JKB: conceptualisation, methodology, software, validation, writing – review & editing, supervision

ZJ: conceptualisation, methodology, investigation, writing – review & editing, supervision

## Data Availability

All data produced in the present work are contained in the manuscript and supplementary materials.

