## Supplementary Figures for "Integrated genomic meta-analysis identifies cell-type-specific signatures and targets in early Alzheimer’s disease"

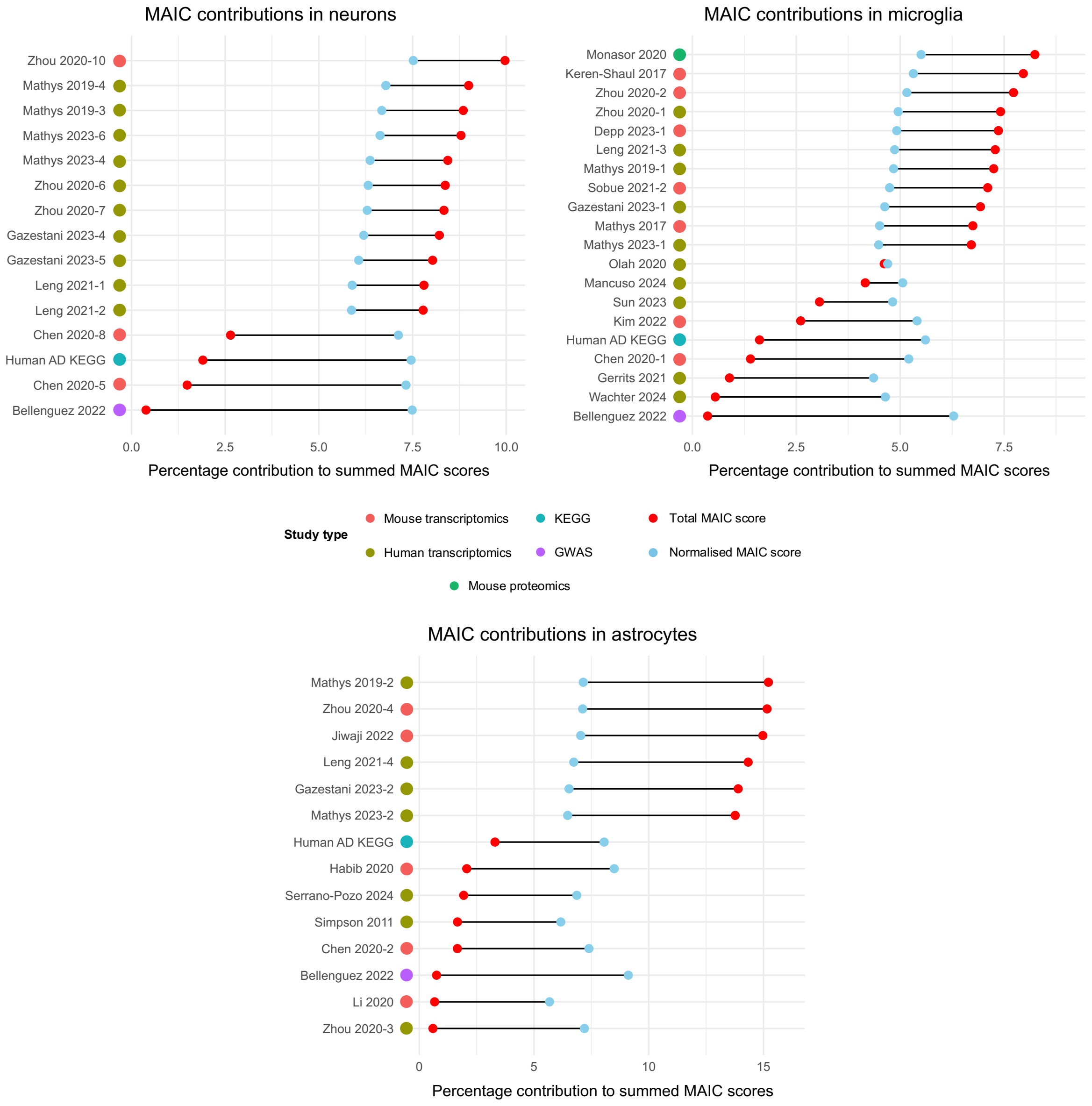


**Supplementary Figure 1:** Percentage contributions of individual studies to MAIC scores in each cell-type. For each study, the MAIC scores of individual genes were summed to obtain a total study-specific MAIC score. This was divided by the sum of MAIC scores across all studies within that cell-type to calculate the study's relative contribution. To account for differences in gene list length, these calculations were repeated after normalising study-specific MAIC scores by the number of genes in their list. Studies are labelled by their data source. KEGG = Kyoto Encyclopaedia of Genes and Genomes, GWAS = genome-wide association study.

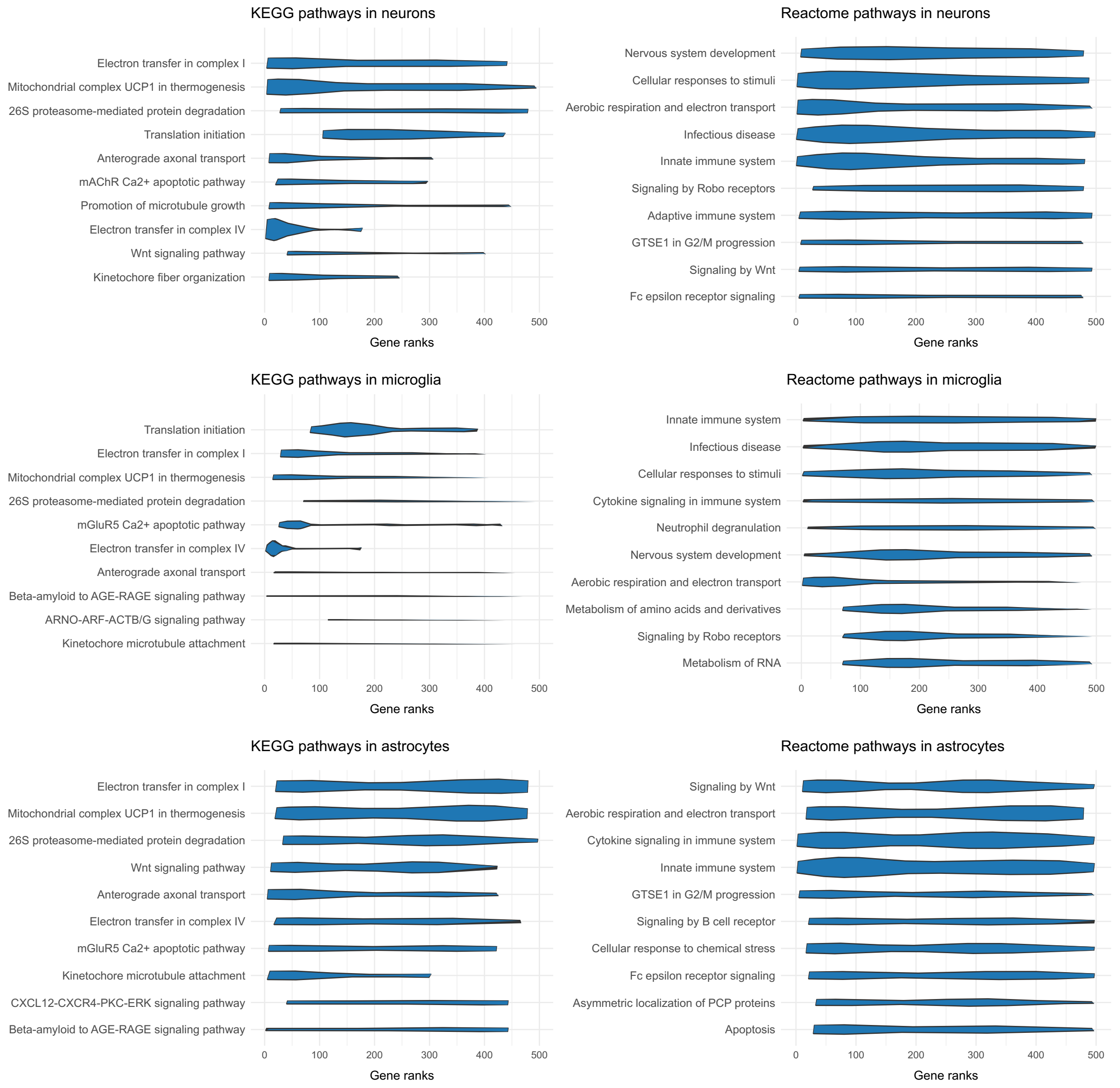


**Supplementary Figure 2:** Top 10 significantly enriched KEGG and Reactome pathways in each cell-type. Repeat occurrences of highly similar pathways in one cell-type were excluded. Violin plots display the rank distributions of genes which appear in each pathway. Pathways are ranked by their false discovery rate (FDR). All pathways have an FDR of < 0.05. KEGG = Kyoto Encyclopaedia of Genes and Genomes.
